# Lower left ventricular ejection time in *MYBPC3* variant carriers with overt or subclinical hypertrophic cardiomyopathy

**DOI:** 10.1101/2024.10.15.24315469

**Authors:** Isabell Yan, Zoe Möhring, Daniel Reichart, Julia Münch, Rixa Woitschach, Paulus Kirchhof, Lucie Carrier, Carolyn Y. Ho, Thomas Eschenhagen, Monica Patten

**Affiliations:** Department of Cardiology, University Heart and Vascular Center Hamburg, University Medical Center Hamburg-Eppendorf, Hamburg, Germany; Cardiovascular Division, Brigham and Women’s Hospital, Boston, USA; Department of Medicine I, University Hospital, Ludwig-Maximilians-University Munich, Germany; Institute for Human Genetics, University Medical Center Hamburg-Eppendorf, Hamburg, Germany; Institute of Experimental Pharmacology and Toxicology, University Medical Center Hamburg-Eppendorf, Hamburg, Germany; German Centre for Cardiovascular Research (DZHK), Partner Site Hamburg/Kiel/Lübeck, Hamburg, Germany

**Keywords:** hypertrophic cardiomyopathy, left ventricular ejection time, genetic variant, *MYBPC3*, *MYH7*

## Abstract

**Aims:** Hypertrophic cardiomyopathy (HCM) is an inherited cardiomyopathy mainly caused by pathogenic variants in *MYBPC3* and *MYH7,* encoding myosin binding protein C3 and myosin heavy chain 7, respectively. These variants can cause increased actin-myosin crossbridge cycling resulting in ventricular hypercontractility. Little is known about genotype-specific differences. Mice lacking *Mybpc3* exhibited reduced left ventricular ejection time (LVET). In this study we tested whether LVET is specifically altered in patients carrying *MYBPC3* variants by retrospective echocardiographic analysis in two genotype-defined HCM cohorts.

**Methods and results:** LVET was measured by echocardiography and adjusted for heart rate (LVET index, LVETI) in 173 patients carrying *MYBPC3* or *MYH7* pathogenic variant. There was a discovery cohort (Hamburg; 46 *MYBPC3,* 31 *MYH7)* and a validation cohort (“Valsartan in Attenuating Disease Evolution in Early Sarcomeric HCM”; 55 *MYBPC3*, 41 *MYH7*). Data were compared with 44 healthy controls from Hamburg. Variant carriers were stratified for overt (G+LVH+) or subclinical left ventricular hypertrophy (G+LVH-). LVETI was lower in *MYBPC3* and higher in *MYH7* G+LVH+ patients than in controls in the discovery, validation and pooled cohorts (pooled: 385 ± 23 ms *MYBPC3*, 436 ± 38 ms *MYH7,* 411 ± 15 ms controls). Similar findings were seen in G+LVH-.

**Conclusion:** The data suggest that variants in *MYBPC3* and *MYH7* result in distinct biophysical consequences, which can be detected by measuring LVETI in patients. The findings may have implications for potential genotype-specific differences in response to therapies targeting sarcomere function.

## Introduction

Hypertrophic cardiomyopathy (HCM) is characterized by thickening of the left ventricular wall, contractile abnormalities, and potentially fatal arrhythmias. It is primarily caused by variants in genes such as *MYBPC3* and *MYH7,* encoding myosin binding protein C3 and myosin heavy chain 7, respectively.^1^ Pathophysiologic consequences are altered sarcomere function leading to prolonged relaxation kinetics, oxidative and metabolic stress, and hyperdynamic contractility.^2–4^

The clinical presentation of patients with HCM carrying *MYH7* or *MYBPC3* variants appears similar by standard imaging techniques.^5^ HCM arising from either disease gene are associated with a broad range of clinical manifestations, from asymptomatic to advanced heart failure and sudden cardiac death. However, the consequences of variants in *MYH7* and *MYBPC3* differ at the molecular level. Pathogenic variants in *MYH7* are mainly missense, resulting in abnormal proteins that interfere with normal sarcomere function.^6^ In contrast, pathogenic *MYBPC3* variants, which account for the majority of genetic HCM cases, are typically truncating, leading to aberrant splicing and reduction of total MYBPC3 protein abundance (haploinsufficiency).^7^ Homozygous *Mybpc3* knock-out mice present with dilated cardiomyopathy and LV hypertrophy,^8^ and reduced left ventricular ejection time (LVET).^9^

LVET reflects the duration of contraction and is a marker of left ventricular contractile function, distinct from peak force, reflected by left ventricular ejection fraction (LVEF). It is not yet used in routine clinical testing but has gained interest as an indicator of inotropic drug action. LVET is also considered a direct measure of stroke volume,^10^ and reduced LVET was shown to be an independent predictor for heart failure and mortality.^11^

Here we aimed to test the hypothesis that reduced LVET may be a specific alteration in *MYBPC3-*related HCM by performing echocardiographic analysis of genotyped cohorts of patients with HCM.

## Methods

### Cohort characteristics

LVET was first retrospectively determined in patients with HCM seen at the outpatient clinic at the University Heart and Vascular Center Hamburg and compared with data from age- and sex-matched healthy controls of the Hamburg cohort from the MOVE Study.^12^ The results were validated in participants of the “Valsartan in Attenuating Disease Evolution in Early Sarcomeric HCM” (VANISH) trial.^13^ In the pooled cohort, 145 patients with HCM had a genetically confirmed pathogenic variant class 4 or 5 and an IVSd (interventricular septal diameter in diastole) ≥ 13 mm, G+LVH+), while 28 patients carrying variant class 4 or 5 had a subclinical phenotype (IVSd < 13 mm, G+LVH-). A cutoff value of IVS = 13 mm was used according to current recommendations for HCM classification.^14^

LVET (defined as the time from opening of the aortic valve to its closure) was measured from transthoracic echocardiography by two independent investigators, who were blinded to genotype. Since LVET is inversely related to heart rate, LVET index (LVETI) was used assuming a linear relationship between LVET and heart rate. The formula was derived on the basis of sex-specific resting regression equations (for female: LVETI = 1.6 x HR + LVET, and for male: LVETI = 1.7 x HR + LVET). Normal values for LVETI are 413 ± 10 ms in males and 418 ± 10 ms in females.^15^

Analyses of genetic variants and LVETI were adjusted for other disease-specific parameters (LVEF, IVSd, left ventricular outflow tract [LVOT] gradient) and for other clinical characteristics (age, sex, concomitant medication use). Analyses were performed in G+LVH+ and G+LVH-groups.

### Statistics

The power calculation assumed an LVETI difference of 12 ms between the control group and patients with HCM mutation carriers, based on a study on patients with diastolic dysfunction.^15,16^ It resulted in a minimum sample size of n=18 and n=36 for single and combined cohorts, respectively, at a desired p-value of 0.05 and a power of 95%.

For continuous variables (LVETI, age, LVEF, IVSd, LVOT gradient at rest, LVOT gradient during exercise), the mean value and standard deviation (SD) were specified; the nominal data (gender, medication intake) were presented using absolute and relative proportions (number and percentage). Analyses were performed comparing *MYH7* and *MYBPC3* variant carriers with either overt or subclinical HCM (G+LVH+, G+LVH-).

Unpaired Student’s t-test was used to compare continuous variables.

A multivariate analysis of variance (MANOVA) was used to identify differences in LVETI in variant carriers and controls with LVETI as the dependent variable, adjusted for gender, age, and atrial fibrillation/sinus rhythm, LVEF, IVSd and heart rate. To analyze the pattern of differences of means between two groups, we used Fisheŕs least significant difference (LSD) method as a post-hoc test to perform the pairwise comparisons.

The stored residuals were visually checked for normal distribution. All analyses were performed with *IBM SPSS Statistics*; a p-value <0.05 was considered statistically significant. Boxplots were generated with GraphPad Prism version 5.

## Results

### Discovery cohort

The discovery cohort comprised 46 patients from Hamburg, Germany with HCM carrying *MYBPC3* variants and 31 with *MYH7* variants (Table). The mean age was 52 years and 47 years, respectively. The mean age of controls was 44 years. The percentage of women was 45%, 64% and 50% in *MYBPC3*, *MYH7* and control groups, respectively. Healthy controls had an LVETI of 411 ± 15 ms, patients with *MYBPC3* variants 395 ± 27 ms (p=0.002 vs. controls), and patients with *MYH7* 423 ± 33 ms (p<0.001 vs. *MYBPC3* and p=0.03 vs. controls).

**Table:** Clinical characteristics of patients with HCM and controls. The patients were separated in groups with overt HCM (G+LVH+, IVSd ≥ 13 mm) or subclinical HCM (G+LVH-, IVSd < 13 mm) and compared to a control cohort from Hamburg, Germany. Abbreviations: *MYH7* myosin heavy chain 7 gene; *MYBPC3* myosin binding protein C3 gene; VANISH,Valsartan in Attenuating Disease Evolution in Early Sarcomeric HCM; LVEF left ventricular ejection fraction; IVSd interventricular septal thickness in diastole; LVOT left ventricular outflow tract; ACE Angiotensin-Converting Enzyme, n number of individuals.

|  | Control (n=44) | MYH7 (n=72) |  |  |  | MYBPC3 (n=101) |  |  |  |
| --- | --- | --- | --- | --- | --- | --- | --- | --- | --- |
|  |  | G+LVH- (n=13) |  | G+LVH+ (n=59) |  | G+LVH- (n=15) |  | G+LVH+ (n=86) |  |
|  | Hamburg (n=44) | Hamburg (n=3) | VANISH (n=10) | Hamburg (n=28) | VANISH (n=31) | Hamburg (n=4) | VANISH (n=11) | Hamburg (n=42) | VANISH (n=44) |
| Age, years, mean (SD); min - max | 44 (12); 21-63 | 26 (3); 23-29 | 27 (7); 19-44 | 47 (15); 18-79 | 30 (8); 18-43 | 34 (13); 24-53 | 21 (4); 18-31 | 52 (17); 20-86 | 30 (8); 18-43 |
| Female sex, n (%) | 22 (50) | 1 (33) | 8 (80) | 18 (64) | 12 (39) | 3 (75) | 4 (36) | 19 (45) | 14 (32) |
| LVEF, %, mean (SD) | 62 (4) | 57 (1) | 59 (2) | 56 (4) | 59 (3) | 57 (1) | 60 (3) | 56 (7) | 60 (3) |
| IVSd, mm, mean (SD) | 9 (3) | 10 (2) | 11 (1) | 22 (5) | 18 (4) | 11 (2) | 11 (2) | 22 (7) | 17 (4) |
| LVOT gradient at rest, mmHg, mean (SD) | <5 | 6 (2) | 5 (1) | 11 (9) | 6 (2) | 6 (1) | 5 (1) | 11 (13) | 7 (6) |
| LVOT gradient max, mmHg, mean (SD) | <5 | 6 (2) | / | 22 (24) | / | 6 (1) | / | 17 (20) | / |
| ACE inhibitor, n (%) | 0 (0) | 1 (33) | 0 (0) | 2 (7) | 0 (0) | 0 (0) | 0 (0) | 7 (17) | 0 (0) |
| Beta blocker, n (%) | 0 (0) | 0 (0) | 0 (0) | 18 (64) | 12 (68) | 0 (0) | 0 (0) | 33 (79) | 8 (34) |
| Calcium channel blockers, n (%) | 0 (0) | 1 (33) | 0 (0) | 2 (7) | 2 (9) | 0 (0) | 0 (0) | 4 (10) | 2 (8) |

Subgroup analyses were performed on G+LVH+ patients (42 *MYBPC3,* 28 *MYH7*) and G+LVH-patients (4 *MYBPC3,* 3 *MYH7*). *MYBPC3* G+LVH+ patients had lower LVETI than both controls (394 ± 25 ms vs. 411 ± 15 ms; p = 0.002) and *MYH7* G+LVH+ patients (437 ± 38 ms (p<0.001 vs. *MYBPC3*; Figure 1). LVETI was also lower in *MYBPC3* G+LVH-than in *MYH7* G+LVH-patients (394 ± 9 ms vs. 415 ± 26 ms, p=0.02; Figure 1).

**Figure 1.**
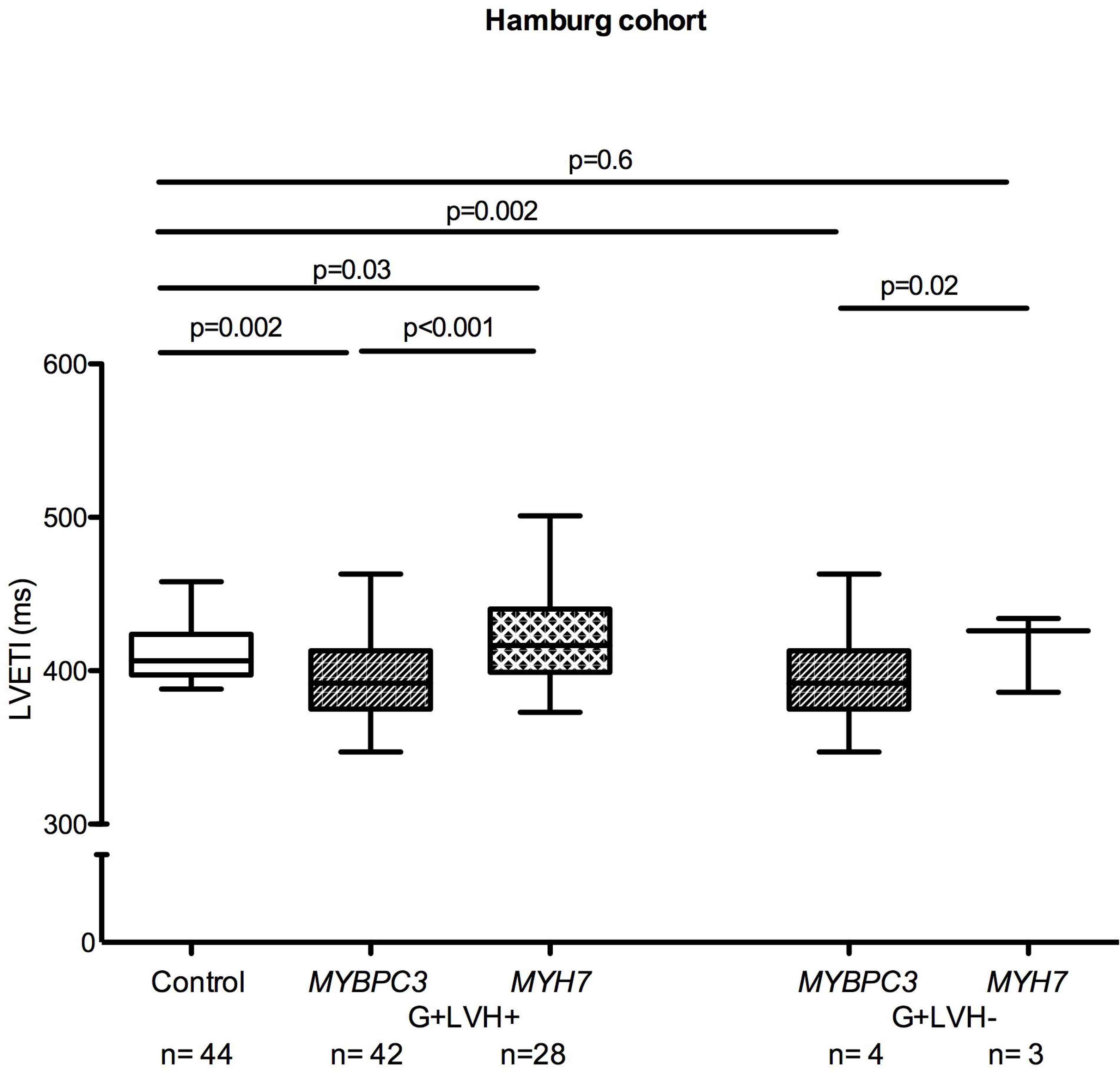
LVETI in healthy controls and HCM patients in the discovery cohort. The patients were separated in groups with overt HCM (G+LVH+, IVSd ≥ 13 mm) or subclinical HCM (G+LVH-, IVSd < 13 mm). The statistical significance was assessed by MANOVA. A linear model was used for comparison between groups, adjusted for age, sex, rhythm, left ventricular ejection time (LVEF), interventricular septum in diastole (IVSd) and heart rate. Data are shown as median and 1.5 IQR. Abbreviations: LVETI left ventricular ejection time index; LVH left ventricular hypertrophy; *MYH7* myosin heavy chain 7 gene; *MYBPC3* myosin binding protein C3 gene; n number of individuals.

### Validation cohort

The validation cohort from the VANISH Trial comprised 55 patients carrying *MYBPC3* variants and 41 patients carrying *MYH7* variants. The mean age was 28 years and 29 years, respectively. The percentage of women was 32% and 39% in *MYBPC3* und *MYH7* groups, respectively. Patients with *MYBPC3* variants had an LVETI of 376 ± 17 ms (p<0.001 vs. controls) and *MYH7* carriers an LVETI of 447 ± 39 ms (p<0.001 vs. *MYBPC3* and control groups.

In the subgroup analysis, LVETI was lower in *MYBPC3* G+LVH+ than in both controls (376 ± 18 ms vs. 411 ± 15 ms; p<0.001) and *MYH7* G+LVH+ (449 ± 39 ms, p<0.001 vs. *MYBPC3*; Figure 2). Similarly, LVETI was lower in *MYBPC3* G+LVH-(377 ± 16 ms) than in *MYH7* G+LVH-(443 ± 40 ms; p<0.001; Figure 2).

**Figure 2.**
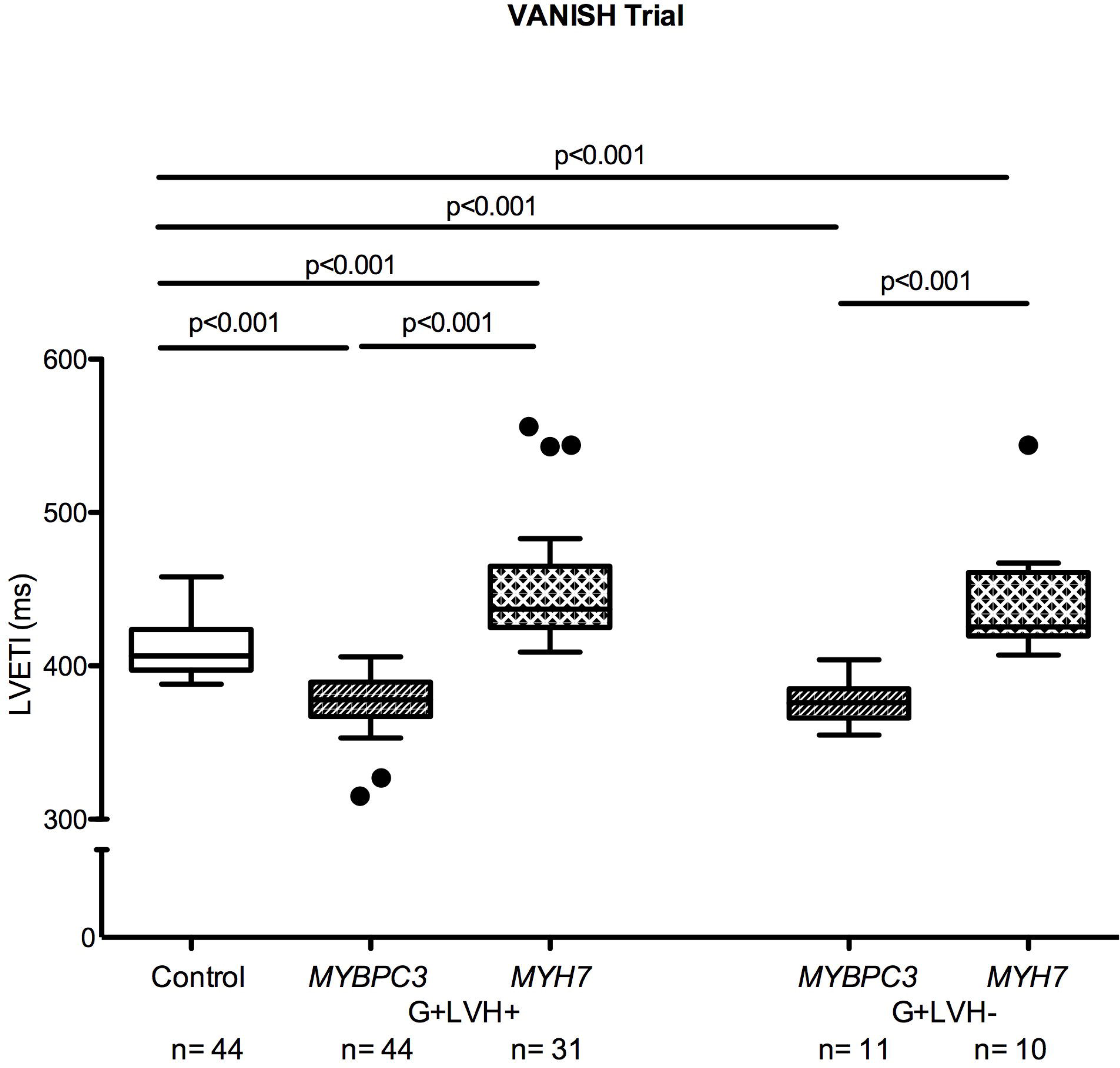
LVETI in healthy controls and HCM patients in the validation cohort. The participants of the VANISH trial were separated in groups with overt HCM (G+LVH+, IVSd ≥ 13 mm) or subclinical HCM (G+LVH-, IVSd < 13 mm). The statistical significance was assessed by MANOVA. A linear model was used for comparison between groups, adjusted for age, sex, rhythm, left ventricular ejection time (LVEF), interventricular septum in diastole (IVSd) and heart rate. Data are shown as median and 1.5 IQR. Abbreviations: LVETI left ventricular ejection time index; LVH left ventricular hypertrophy; *MYH7* myosin heavy chain 7 gene; *MYBPC3* myosin binding protein C3 gene; n number of individuals.

### Pooled cohort

Combining the discovery and validation cohorts, patients with *MYBPC3* variants (n=101) showed the shortest LVETI (385 ± 23 ms; p<0.001 vs. control), and *MYH7* carriers (n=72) the longest LVETI (436 ± 38 ms; p<0.001 vs. *MYBPC3* and control groups).

In the subgroup analysis, LVETI was lower in *MYBPC3* G+LVH+ (n=86, 385 ± 24 ms) than in both *MYH7* G+LVH+ patients (n=59, 437 ± 38 ms; p<0.001) and controls (n=44, 411 ± 15 ms; p<0.001 vs. both; Figure 3). LVETI was also lower in *MYBPC3* G+LVH-(n=15, 381 ± 16 ms) than in *MYH7* G+LVH-(n=13, 436 ± 39 ms, p<0.001) and controls (p=0.003 vs. *MYBPC3*, p=0.004 vs. *MYH7*; Figure 3).

**Figure 3.**
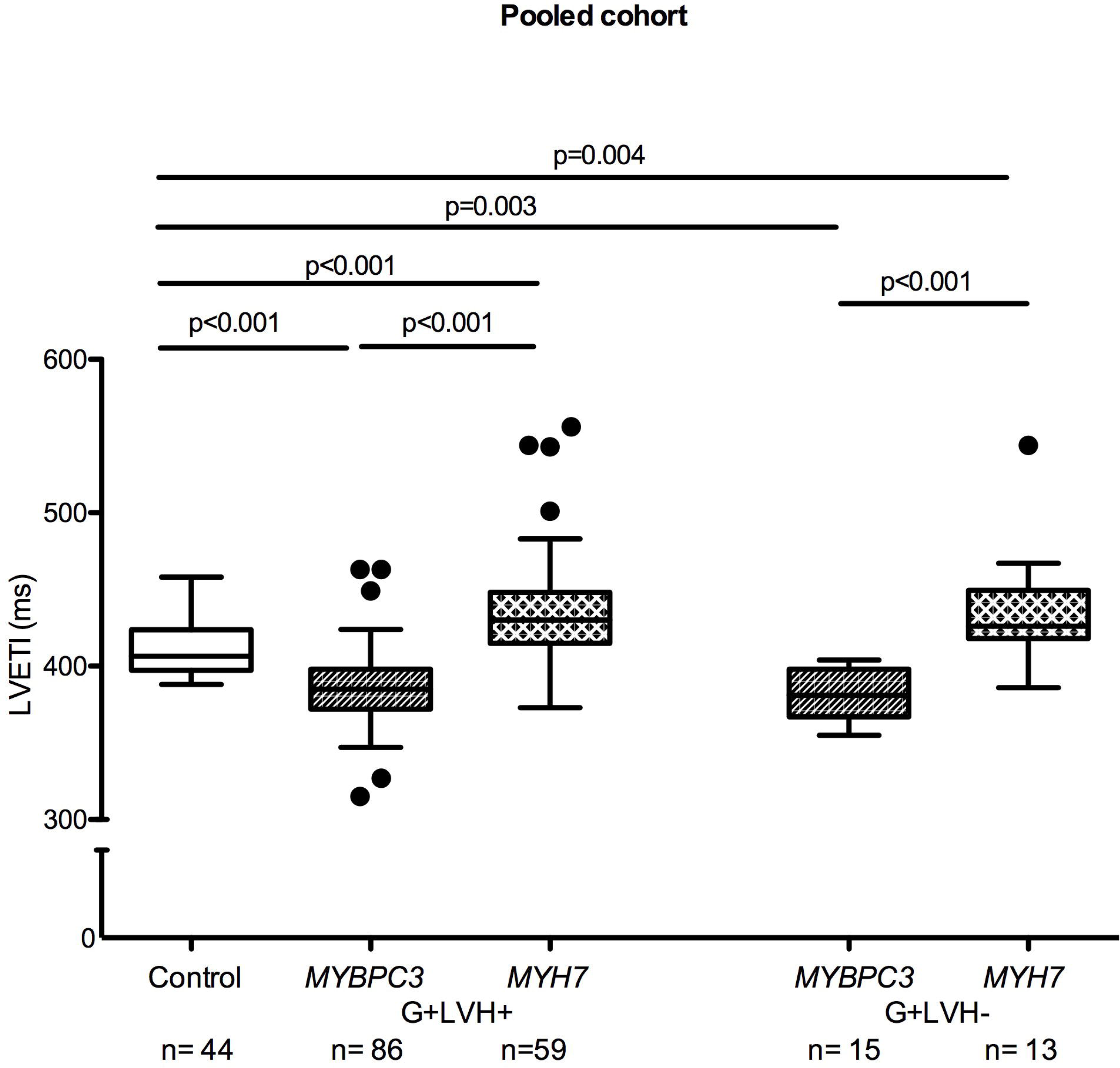
LVETI in healthy controls and HCM patients in the pooled cohort. The patients were separated in groups with overt HCM (G+LVH+, IVSd ≥ 13 mm) or subclinical HCM (G+LVH-, IVSd < 13 mm). The statistical significance was assessed by MANOVA. A linear model was used for comparison between groups, adjusted for age, sex, rhythm, left ventricular ejection time (LVEF), interventricular septum in diastole (IVSd) and heart rate. Data are shown as median and 1.5 IQR. Abbreviations: LVETI left ventricular ejection time index; LVH left ventricular hypertrophy; *MYH7* myosin heavy chain 7 gene; *MYBPC3* myosin binding protein C3 gene; n number of individuals.

Other disease specific parameters, such as LVEF, IVSd, and LVOT gradient at rest did not reveal statistically significant differences between *MYBPC3* and *MYH7* patients with HCM (Table).

## Discussion

This study demonstrates distinct genotype-related differences in the duration of contraction in patients with HCM. LVETI was significantly lower in individuals with pathogenic *MYBPC3* variants than in both individuals with *MYH7* variants and healthy controls. The differences in mean LVETI between *MYBPC3* carriers and controls (−26 ms) and between *MYH7* carriers and controls (+25 ms), respectively, are larger than the reported difference between patients with diastolic dysfunction and controls (−12 ms).^15,16^ Similarly, they were similar or larger than those reported for patients with mildly, moderately and severely reduced ejection fraction (−4 ms, −8 ms, −28 ms) compared to healthy controls,^17^ arguing for their pathophysiological relevance. The differences in LVETI were not only observed in variant carriers with clinically overt HCM (G+LVH+), but also in G+LVH-, suggesting that alterations in ejection time are a primary manifestation of *MYBPC3* and *MYH7* genetic variations itself. This and the opposing direction of changes in LVETI in *MYBPC3* and *MYH7* variant carriers indicate that they are not a general consequence or feature of HCM.

LVETI was lower in the *MYBPC3* than in the *MYH7* group despite similar degrees of cardiac hypertrophy, outflow tract obstruction and LVEF, indicating that *MYBPC3* variants may be associated with a specific deficit in the late contraction/early relaxation phase. These findings support prior mouse and tissue studies.^1,2,9,18^ Genetically induced homozygous reduction or loss of *MYBPC3* in mice led to reduced LVET,^9,18^ and human septal myectomy samples showed higher rates of the fast-exponential phase of relaxation.^19^ Additionally, shorter contraction time was seen in engineered heart tissues from *Mybpc3*-targeted knock-in mice.^20,21^ These data support the hypothesis that *MYBPC3* not only facilitates diastolic relaxation by promoting a super-relaxed state of myosin, but also stabilizes systole by binding to the thin filament.^9^ Thus, reduction or loss of *MYBPC3* may reduce the ability of the heart to maintain systolic ejection. In support of this hypothesis, Beltrami et al. recently showed in a retrospective analysis that *MYBPC3* variant carriers were more likely to develop systolic dysfunction over time than *MYH7* carriers.^22^ This data is somewhat at odds with earlier data suggesting a better survival prognosis of *MYBPC3* than *MYH7* carriers^23^ and certainly requires validation. But it is in support of preclinical^9,18,20,21^ and the present data, pointing at a specific systolic deficit conferred by *MYBPC3* variants.

In contrast to routinely measured echocardiographic parameters such as LVEF and IVSd thickness, LVET has not yet found its way into routine clinical practice. However, it is increasingly recognized as an important metric of cardiac function and drug response in patients with heart failure and cardiomyopathies.^10^ As disease-specific medications for HCM are beginning to enter clinical practice, including cardiac myosin inhibitors developed to slow cycling rate,^24,25^ these findings emphasize gene-specific differences in contractile function and may impact the response to therapies that target different aspects of sarcomere function. Although further study in larger cohorts is required, our results also suggest that contractile abnormalities in HCM may be driven by the genetic variant and not just the hypertrophic remodeling.

### Conclusion

In conclusion, our study provides evidence for distinctive decreased duration of contraction in patients with HCM carrying *MYBPC3* variants. These data suggest that the reduction in ejection time may be a specific manifestation of *MYBPC3* genetic defect, which can be visualized even before clinically overt disease develops. These findings may have implications for the natural history of disease as well as response to targeted therapies.

## Data Availability

All data produced in the present work are contained in the manuscript.

## Acknowledgements

The authors thank the VANISH investigators and participants for their contribution: Murilo O. Antunes Edmundo Arteaga, Jose E. Krieger, Luciana Sacilotto (Laboratory of Genetics and Molecular Cardiology, Heart Institute, University of São Paulo Medical School, São Paulo Brazil), Euan Ashley (Division of Cardiovascular Medicine, Stanford University School of Medicine, Stanford, CA, USA), Kimberley Y. Lin (Children’s Hospital of Philadelphia, Philadelphia, PA, USA), E. Kevin Hall (Yale University School of Medicine, New Haven, CT, USA), Lubna Choudhury (Feinberg School of Medicine, Bluhm Cardiovascular Institute, Chicago, IL, USA), Elfriede Pahl, Philip Thrush (Ann & Robert H. Lurie Children’s Hospital of Chicago, Chicago, IL, USA), Harry M. Lever (Cleveland Clinic Foundation, Cleveland, OH, USA), Renee Margossian (Department of Cardiology, Boston Children’s Hospital, Boston, MA, USA), Lee Benson (Toronto Hospital for Sick Children, Toronto, ON, Canada), Allison C. Cirino (Cardiovascular Division, Brigham and Women’s Hospital and Harvard Medical School, Boston, MA, USA), Eugene Braunwald, Akshay S. Desai, Calum A. MacRay, E. John Orav, Scott D. Salomon (Brigham and Women’s Hospital and Harvard Medical School, Boston, MA, USA), Christine E. Seidman (Brigham and Women’s Hospital and Harvard Medical School, Howard Hughes Medical Institute, Boston, MA, USA), Kristin M. Burns (Division of Cardiovascular Sciences, National Heart, Lung, and Blood Institute, National Institutes of Health, Bethesda, MD, USA), John J.V. McMurray (British Heart Foundation Cardiovascular Research Centre, University of Glasgow, Glasgow, Scotland, UK), Gregory D. Lewis (Cardiac Unit, Massachusetts General Hospital and Harvard Medical School, Boston, MA, USA).

## Fundings

P.K. is partially supported by grants from the European Union (AFFECT-AF (847770) and MAESTRIA (965286)), the British Heart Foundation (PG/20/22/35093 & AA/18/2/34218), the German Centre for Cardiovascular Research (DZHK, FKZ 81X2800182, 81Z0710116, 81Z0710110), the German Ministry of Research Education (BMBF) and the German Research Foundation (Ki 509167694).

L.C. is supported by grants from the Leducq Foundation (20CVD01), the German Centre for Cardiovascular Research (DZHK) and the German Ministry of Research Education (BMBF).

C.Y.H. is funded by the National Institutes of Health (USA) and receives unrestricted research funding from Bristol Myers Squibb, Pfizer, Cytokinetics, Biomarin, Tenaya, viz.AI, Lexicon.

The VANISH trial was funded by the National Institutes of Health (NIH/NHLBI P50HL112349).

T.E. is supported by grants from the German Research Foundation (DFG Es 88/16-1, Es 88/17-1) and the European Union’s Horizon 2020 research and innovation program (874764).

M.P. receives funding from Takeda/Shire.

## Conflict of interest

P.K. received research support for basic, translational, and clinical research projects from several drug and device companies active in atrial fibrillation and has received honoraria from several companies in the past, but not in the last five years. PK is listed as inventor on two issued patents held by University of Hamburg (Atrial Fibrillation Therapy WO 2015140571, Markers for Atrial Fibrillation WO 2016012783).

L.C. and T.E. are advisors and shareholders of DiNAQOR AG developing a *MYBPC3*-based gene therapy for HCM.

C.Y.H. receives consulting honoraria from Bristol Myers Squibb, Pfizer, Cytokinetics, Biomarin, Tenaya, viz.AI, Lexicon.

M.P. receives consulting honoraria from Bristol Myers Squibb, Cytokinetics, Sanofi, Alnylam and Pfizer.

I.Y., Z.M., D.R., J.M., and R.W have nothing to disclose.

## Notes

### Author Declarations

-Ethikkommission der Aerztekammer Hamburg, Germany gave approval -approval were granted by consensus of the executive committee from the VANISH Trail

